# Stakeholders’ perception of possible integration pathways for eye health into school health programme in Zanzibar: a qualitative health system research

**DOI:** 10.1101/2021.07.25.21260645

**Authors:** Ving Fai Chan, Elodie Yard, Eden Mashayo, Damaris Mulewa, Lesley Drake, Fatma Omar

**Author notes:** Corresponding Author: Dr Ving Fai Chan, Centre of Public Health, School of Medicine, Dentistry and Biomedical Medicine, Institute of Clinical Sciences, Block B. Royal Victoria Hospital. Belfast. BT12 6BA, Queen’s University Belfast. was affiliation when the study was undertaken. This paper has not been published nor submitted simultaneously for publication elsewhere. We have not published, posted or summited any related papers from the study.

## Abstract

**Purpose:** To scope the potential for eye health programme to be integrated into Zanzibar School Health programme, through the lenses of stakeholders

**Methods:** Embedded into an operational research project integrating eye and School health, we elicited responses from 83 participants, purposefully selected from the Ministry of Health (n=7), Ministry of Education and Vocational Training (n=7), hospitals/eye centres (n=5), master trainers (4) and schools (n=60) participated in in-depth interviews. Their responses were analysed and grouped into four pre-determined themes of Human Resource Training, Resources Mobilisation, Acceptability, and Leadership and Governance. Quotations are presented to illustrate the findings.

**Results:** In line with the four research themes, i) The integrated school eye health programme training was satisfactory, with room for improvement, ii) Child eye health screening conducted by teachers was well-received, with concerns and suggestions to improve its effectiveness and efficiency, iii) Integration of eye health into the school health programme is perceived as a good initiative, but to increase referral, service uptake and spectacle usage, primary care units must be equipped, and eye health awareness needs to be improved, iv) Departmental roles, resources, gaps and synergies to ensure eye health is successfully integrated into the school health programme.

**Conclusion:** The concept of integrated school eye health delivery is generally well-received by beneficiaries and stakeholders within an operational research project in Zanzibar, with the caveat that investment is required for effective referral and update.

## Introduction

Poor health is associated with 200 to 500 million lost school days annually in low-income countries.^1^ Some of the most common health conditions, including poor vision, negatively affect educational performance among school-age children.^2^ Furthermore, poor child eye health is closely associated with low self-esteem, poor cognitive abilities,^3,4^ reduced quality of life, and reduced future economic productivity.^5,6^ However, the prevalence of childhood blindness is high – with an estimated 8,500 to 10,000 blind children live in East Africa^7^ and Zanzibar alone, and 42% of the children in rural communities who need it did not have a pair of glasses.^8^

Recognising the importance of good vision, many countries have made eye health an essential part of school health programmes. In low- and middle-income countries, these programmes are usually implemented vertically with non-governmental organisations (NGO) support. Given the short-term funding for these programmes, these vertical approaches do not strengthen the local health systems and limit long-term programme impacts. Despite the great need for integration, the evidence of integrating eye health into mainstream school health programmes to ensure effectiveness and efficiency is currently weak.^9,10^

In Zanzibar, child eye health services, such as basic eye examination, eye drops distribution and refraction services, are provided at all levels of health care (district, regional and national). To improve services uptake at the service points, ad hoc-school eye health programmes were conducted, often under-resourced. For example, from 2012– 2016, eye health outreach programmes were conducted with NGO’s support and, despite achieving high screening and treatment rates, the programme had to come to an end when the funding ceased (Zanzibar Eye Health Project, 2017, unpublished).

Recognising the need for practical intervention by NGOs and ensuring the school eye health programme’s sustainability, the Zanzibar Government seeks to integrate eye health into the school health programme. While the current school health programme focuses on water and sanitation and food and nutrition, the Revolutionary Government of Zanzibar identified that public health practices and access to disability-related services should be improved (Eye Health Strategic Plan 2018 – 2022, unpublished). Hence, the Government aims to integrate school eye health with the school health programme. Aligning with the aim of Focusing Resources on Effective School Health, A FRESH Approach for Achieving Education for All *“to implement school-based health programmes in efficient, realistic and results-oriented ways*”.^11^

An implementation research to compare the performance of an integrated and vertical school health eye programme was collaboratively conducted by the Ministry of Health and local key stakeholders, an eyecare non-governmental organisation and a child health and nutrition research and technical assistance group from April to October 2017. It was found that the integrated model achieved 96% screening coverage, the cost per child screened was only $1.23, and the cost per child identified as having an eye problem in the integrated model was only half that of the vertical model ($24.76 vs $51.75).^12^ Subsequently, a series of in-depth interviews using a systematic approach was conducted. The objective of the research was to discuss the implementation stakeholders’(Ministry of Health, Ministry of Education, hospitals/eye centres, master trainers and school representatives) views on an integrated school eye health programme and their suggestions on how to integrate it into the mainstream school health programme fully.

## Methodology

This study’s data was collected as part of the larger implementation research aimed at building the evidence base for an effective school eye health intervention in Zanzibar (the parent study). The study consisted of a quantitative study to compare the performance and costs of an integrated and a vertical school health eye programme;^12^ and a qualitative study to obtain the partners’ views on the future implementation of the integrated school eye health programme and how to realise this. This article focuses on the qualitative study.

The study protocol was approved by the Zanzibar Medical Research and Ethics Committee (ZAMREC/0001/January/17). Written consent was obtained from the respondents before they participated in the study. According to the Consolidated Criteria for Reporting Qualitative Research (COREQ), this paper was prepared: a 32-item checklist for interviews and focus groups.

Two qualitative interviewers, each with social science backgrounds (a male and a female), were trained to conduct the in-depth interviews. Both researchers have more than ten years of experience implementing school health projects in Tanzania, including integrating health initiatives into the mainstream health system.

### Interview respondents- sample and composition

The 83 participants, purposefully selected from the Ministry of Health (n=7), Ministry of Education and Vocational Training (n=7), hospitals/eye centres (n=5), master trainers (4) and the 19 schools (n=60), participated in the study. These participants were selected to provide rich and diverse information and practical recommendations on integrating eye health into the school health programme in Zanzibar. The interviews were conducted at the participants’ offices with no third-party present to ensure that they were comfortable giving their responses. Table 1 shows the composition of the respondents.

**Table 1:** The composition and numbers of the respondents who participated in the in-depth interviews

| **Respondents**<br>**(number)** | **Composition** |
| --- | --- |
| Ministry of Health interviewees (n=7) | Director-General, Director of Planning, Policy and Research. Director of Preventive Service and Health Promotion, District Medical Officers from Micheweni, Mkoani, North A and Unguja districts |
| Ministry of Education interviewees (n=7) | Director, Department of Policy and Planning, Deputy Principal Secretary, Director for Pre-primary and Primary Education, District Education Officers from Micheweni, Mkoani North A and Unguja |
| Hospitals/eye centres interviewees (n=5) | Three optometrists from Mjini Magharibi, one optometrist from Chake Chake District Hospital and one optometrist from Mnazi Mmoja Hospital |
| Master trainers (n=4) | Two ophthalmic clinical officers and two Home Grown School Feeding Programme officers |
| School interviewees (n=60) | Twenty headteachers and forty school health teachers |

### Data collection

The interviews were conducted using an in-depth interview guide designed in discussions with different local implementing stakeholders and tested in a smaller group to ensure the guide’s content and wording were appropriate. The interview guide is included in a supplementary appendix.

We designed the study by asking the overarching question “How can we integrate eye health into the existing school health programme?” The four specific questions that we wanted to answer were decided with the stakeholders. These were:

- Human resources training - What do they think about the eye health training conducted for the teachers?
- Acceptability - What stakeholders’ acceptability of an integrated school eye health programme that utilises teachers to conduct eye health screening?
- Leadership and governance - How best can the integrated school eye health programme be managed and implemented?
- Mobilisation of resources - What roles do different departments play, and how can we share resources with health programmes with similar interests in eye health?

The interviews took 45 to 60 minutes and were audio-recorded with field notes made during the interviews. The interviews were not repeated because it was challenging to schedule interviews with our respondents who had busy working schedules. Instead, debriefing sessions were conducted with the respondents after the interview to make corrections or add additional comments to the notes.

### Data analysis and reporting

Each interview was transcribed verbatim, comprehensively reviewed, and coded by two data coders (RK and MM). An MS Excel database was created to capture the meaning units and display the systematic relationships between coded texts. The data coders linked the meaning units from the transcripts to similar statements across interviews. To explore the data and conceptualise the findings, related ideas across the interviews were located by bringing together strands of data. Subsequently, while referring to analytic framework of WHO Health System Building Blocks,^13^ the data coders generated the themes and consolidated them into Human Resource Training, Mobilisation of Resources, Acceptability, and Leadership and Governance. Quotations are presented to illustrate the findings. As the number of respondents in some categories was small, it was impossible to anonymise the respondents’ identity. Hence, we assigned the respondents from the different categories into i) the Ministry of Health as MOH 1 to 7; ii) the Ministry of Education and Vocational Training as MOEVT 1 to 7; iii) hospital optometrists as Optom 1 to 5; iv) master trainers as MT 1 to 4; v) headteachers as HT 1 to 20; vi) teachers as TCH 1 to 40. The themes and example quotes from the qualitative interviews are shown in Table 2.

**Table 2:** Themes and example quotes from the qualitative interviews

| Theme | Quote (Representative number) |
| --- | --- |
| Human resources training | 1. The time was short, and a lot had to be covered within a short time. It was quite a challenge for teachers to catch up with the speed and content of the training. (MT 4) |
|  | 2. Consider teacher's age when selecting teachers to be trained, increase training time, conduct supportive supervision, provide incentives for teachers during implementation, train teachers to train other teachers when they return to school. (MT 2) |
|  | 3. This is a good initiative, helps to get early identification of vision impairment in children; however, there is a need to rectify some of the things in the manual. For example, the conjunctiva may somehow be brown, but that does not necessarily mean that a child is sick. So perhaps adjust the section which said white must be white. (MT3) |
|  | 4. Very participatory. Teachers understood through a combination of approaches employed such as lecture, group discussion, plenary presentations and practical exercises on vision screening of peers. (MOEVT 2) |
|  | 5. Separating it will be better to avoid confusion. Each category is to be trained separately. (MOH 1) |
| Acceptability | 6. This is a good initiative. Education goes in line with the ability to see well and not having vision problems. If you cannot see well, it affects your learning process. It is a good way to get early identification of childhood blindness and vision impairment. (MT 1) |
|  | 7. Yes! Because it simplifies work for the optometrist and also increases coverage of eye screening where the optometrist could not manage to reach. We have specialised equipment for eye care, and they can be thoroughly examined (once the children are referred). (Optom 3) |
|  | 8. It went well. They succeeded to identify the pupils who suffer from eye health problems. (The children were) referred to the hospitals. Others did not attend (the hospitals). (TCH 8) |
|  | 9. Some referred pupils did not have any visual problems. They did not necessarily need any referral. Some children were emmetropic, but they (the teachers) fail to take accurate visual acuity. (Optom 1) |
| Leadership and governance | 10. Yes, but this has to be strongly accepted by the policymakers, and there should be proof of the need/problem existing and which clearly indicates that it is worth investing in. (MOH 4) |
| Mobilisation of resources | 11. There is great success in integration. We have done that, and we have seen the results. Unity is strength. We have supported each other in terms of staff and other resources in the integration of Reproductive health and other health interventions. (MOH 1) |
|  | 12. In future, they should book appointments so that when they come, they may not miss me or find that the waiting queue is so long, and this is unbearable for children to wait for so long. School health teachers should have the contact details of the optometrist and enquire about her availability. I am the only optometrist available in the district, and I have many assignments and may at times be out for eye camps etc. The optometrists and all other project implementers must be involved in all stages of the project for smooth operations (especially training). (Optom 5) |
|  | 13. There is no budget for integrated School Eye Health Programmes at the district level, but there is at the Regional Government level. A decision to decentralise the three sectors of agriculture, health (basic health at the health centres) and education (nursery and primary), has been made. (MOEVT 7) |
|  | 14. The best way we can combine efforts with eye health is by having a strategic plan specific for eye health and have a road map that will lead to |
|  | different interventions in eye care, then identify financial resources and how they will be utilised. According to me, primary eye care units must first be strengthened, equipped with enough staff because that is the first referral point from schools. But all this will be achieved if we have a specific road map for eye care. (MOH 1) |
|  | 15. The primary care units have so much potential, but skilled staff specific in eye care lacks primary healthcare units. There is also no adequate eye care equipment in the primary care units. (MOH 5) |
|  | 16. Lack of awareness of both school children and parents, low supervision at school level and lack of collaboration between parents and teachers. (TCH 39) |
|  | 17. Stigmatised by friends/schoolmates, the unwillingness of the pupil, parental support lacking. (HT 14) |

## Results

### Human resources training: The integrated school eye health programme training was satisfactory, with room for improvement

Most of the respondents (n=52) felt that the length of the time allocated for the training (one day for the vertical model and two days for the integrated model) was insufficient. The trainers felt that there was not enough time to cover the training’s content, especially for the integrated model. (Quote 1) Even though the trainers and teachers were satisfied with the training, they suggested increasing training days and focusing on teacher selection, supervision, and incentives to improve training outcomes. (Quote 2)

Almost all the teachers felt that the training materials have enough information. The trainers also commented that the training materials were very clear and could be easily understood. Both trainers and teachers responded that the training, which consisted of group discussions, class participation and lectures, was delivered in a participatory and inclusive manner. (Quote 4) While all the teachers were satisfied with the training, the trainers and the Ministry of Health respondents felt that training for different topics should be conducted separately to avoid confusion, overburdening the teachers and time constraints. (Quote 5)

### Acceptability: Child eye health screening conducted by teachers was well-received, with concerns and suggestions to improve its effectiveness and efficiency

Most of the respondents indicated that eye health screening conducted by teachers is a good initiative and that it worked well. The teachers felt that their contribution to identifying and managing children with eye health problems was recognised. (Quote 6) The teachers also felt that having them conduct eye health screening for children is a practical approach as they spend a lot of time with the students in the schools. The optometrists further commented that the approach simplifies their work and increases screening coverage. (Quote 7) The teachers mentioned strongly that screening high numbers of students interfered with teaching schedules. Furthermore, some children were afraid to be screened and children who failed the eye health screening and did not go to the hospital for further management. (Quote 8) Optometrists further highlighted the issue of teachers referring to children without eye problems to the hospital. (Quote 9)

### Leadership and governance: Departmental roles, resources, gaps and synergies to ensure eye health is successfully integrated into the school health programme

The departmental roles and responsible activities in implementing the school eye health programme in Zanzibar are shown in Table 3. The primary resources suggested maintaining the integrated school eye health programme were the government budget, development partners (community-based organisations, faith-based organisations, non-governmental organisations), community health workers and health centres. It was also pointed out that funding, albeit limited due to competing health priorities, exists in the health care budget to implement and maintain the integrated school eye health programme.

**Table 3:** Perceived departmental roles and responsible activities in school eye health implementation

| Department | Roles | Responsible activities |
| --- | --- | --- |
| **Ministry of Health** | In-charge of primary health care, tertiary health care (primary eye care based in the community); outreaches/camps in schools; monitoring, health promotion and education | School health monitoring programmes in schools |
| **Ministry of Education and Vocational Training** | Design and coordinate support for school children with health problems; supervision of schools involved in the school health programme | Supervision of teachers to implement the school health programme effectively; training of teachers on school health |
| **Preventative Service and Health Promotion Unit** | Coordinate health promotion and education activities, and service provision | School health boards to initiate and support health promotion activities within schools |
| **District Health** | Implement health promotion activities; primary health care, monitoring, school hygiene and immunisation programme | Create awareness of and monitor health promotion activities |
| **District Education** | Supervise and monitor the school health programme at the district level | Training of school teachers and community members; setting up management information systems; resource mobilisation |

Access to the funding depends on the ability to show that an integrated school eye health programme is necessary and cost-effective. (Quote 10) The Ministries’ respondents identified the synergies and gaps between the stakeholders, and shown in Table 4.

**Table 4:** Synergies and gaps identified by Ministries’ respondents in school eye health implementation

| Synergies | Gaps |
| --- | --- |
| ☐ Identifying areas highly affected by eye health problems<br>☐ Planning, implementing and monitoring activities, and addressing operational challenges<br>☐ Conducting teachers training<br>☐ Implementing health promotion activities, which includes community awareness programmes | ☐ School eye health is not mainstreamed; hence it is not budgeted for.<br>☐ District authorities are not involved in planning, and thus there is a lack of proper coordination between regional and district level |

### Mobilisation of resources: Integration of eye health into the school health programme is perceived as a good initiative, but to increase referral, service uptake and spectacle usage, primary care units must be equipped, and eye health awareness needs to be improved

Respondents from the Ministries of Health and Education saw the advantage of integration as the efficient use of resources. They highlighted that unity and coordination are key to successful integration. (Quote 11) The optometrists and the Ministry of Health respondents believed that integrating eye health into the school health programme could be achieved if coordination and clinic space challenges are overcome. (Quote 12) A further major challenge foreseen by the Ministries of Health and Education is the inadequate budget allocation for the school eye health programme. (Quote 13) The Ministry of Health recognises that integrating eye health into the school health programme requires a clear roadmap and resources for implementation. However, identifying and allocating human resources and financial resources was also emphasised as the main challenge in the process. (Quote 14) The respondents felt that district primary care units could be service points for children to access eye health as vision centres do not exist in all districts. However, these primary care units need to be well-equipped, and the staff must be upskilled to handle eye-related issues. (Quote 15) The majority of the respondents expressed their concerns regarding the children’s low spectacle wear. They stated that parents do not encourage their children to wear their spectacles. (Quote 16) Their peers might also tease the children due to the low awareness of eye health’s importance in general. (Quote 17)

## Discussions

While stakeholder engagement is a cornerstone for any health programme integration, there is very little published evidence on this topic. We attempted to methodologically understand how we could integrate eye health into the school health programme in Zanzibar using a health system research approach, which generated positive and insightful results.

### Human resource training

In general, the respondents were satisfied with the training and the training material. To ensure the programme’s effectiveness and efficiency, they suggested more teachers should be trained, with refresher training conducted at regular intervals to maintain and improve the screening quality. Where human resource training was ineffective, there is evidence to show that it was due to inadequate content and quality of training and a lack of support to implement the eye health skills learned.^14–18^ We also recommend that further supervision, ongoing motivation and support is provided

While the teachers in our programme were willing to learn the additional eye health screening skills, caution must be taken to ensure teachers understand that eye health is part of child health so that performing eye health screening is not perceived as an ‘extra’ duty. Hence, it is also critical that the stakeholders understand that the aim is not to train teachers to become eye specialists but to detect children with eye health problems accurately and refer them sufficiently early.^14,19^

### Acceptability

The respondents agreed that it is essential to train teachers in eye health screening, given the high teacher-student contact time. This teacher-led screening approach aligns with Tanzania’s National Eye Health Strategic Plan 2018-2022 and the Integrated People-centred Eye Care (IPCEC) strategy,^20^ to empower and engage people communities in providing eye care to children. Empowering communities has been effective in improving early disease detection and timely intervention, and improving compliance. Furthermore, successful task-sharing by extending responsibilities to lay personnel has shown to increase programme effectiveness.^21^

### Leadership and governance

Published examples of leadership and governance in policy setting and implementation, leading to quality care, are limited in primary eye care.^22^ The first step in integrating eye care is health system planning to create and enable an optimal environment for integration.^20^ This can only be achieved through commitment, good leadership and governance. Hence, the commitment shown by the Zanzibari government and stakeholders is a positive catalyst for integrating eye health in the school health programme.

A sudden increase in patients attending the vision centres following teachers’ referrals was a major challenge. Planning projections used before the pilot proved to be too conservative as the vision centres could not cope with the sudden surge in patient loads. Our pilot provided valuable information for realistic planning to ensure the provision of high-quality care.^23,24^

### Mobilisation of resources

There is no specific budget dedicated to the school eye health programme because it is not a national school health programme component. This a common challenge faced in LMICs.^25^ While the government can allocate resources for school eye health, continuous advocacy must ensure its integration with the national school health programme.

The second and third strategies of IPCEC^20^ emphasises that eye care should be reoriented towards prioritising services delivered at the primary and community level. The aim is for those families who live further away from the vision centres not to be deprived of access to services. It is encouraging that the respondents suggested providing eye care services at the primary health units, which are closer to the communities than the vision centres and private sector optical outlets.

The respondents’ repeated suggestion was to improve the existing eye health education strategy to increase spectacle usage and compliance. To date, there was a minimal investment in child eye health promotion in Zanzibar. However, this is a worldwide phenomenon where attention is focused on treatment. However, health promotion activities have shown to be effective in improving eye health knowledge and awareness in the community, among the older population and those with diabetes, and increasing uptake of eye services in Bangaladesh.^26^

Following the study’s completion, multiple stakeholder discussions have been held with local government ministries, non-governmental organisations, local stakeholders and beneficiaries, leading to the formation of a child eye health forum. Furthermore, the development of an innovative arts-based child eye health education strategy is underway. These initiatives aim to successfully integrate school eye health into the national school health programme for inclusion in the National Health Policy.

In conclusion, the integrated model of school eye health delivery was well-received by the local implementing stakeholders. The main suggestions were:

- improve teachers eye health screening training
- advocate for school eye health integration with the national school health programme and secure specific government budget allocation
- consolidate resources through inter-sectoral collaboration
- offer child eye services at facilities closer to families (such as primary health units)
- improve existing eye health education to increase spectacle compliance and wear

## Data Availability

Data will be made available on at the journal repository.

## Acknowledgement

We thank Mr Kalam Abushir for his commitment to coordinating the project and Ms Rebecca Kasika and Mr Mattias Miti for data collection. Mr Hasan Minto (deceased) for his guidance in the research process.

