## Supplementary file for "Stakeholders’ perception of possible integration pathways for eye health into school health programme in Zanzibar: a qualitative health system research"

| **Research questions** | **Domains** | **Interview Questions** |
| --- | --- | --- |
| How can we integrate eye health into the existing School Health Programme? | **1. General questions relating to the individuals/ departmental role in the programme.** | a. What was your role in the integrated programme? Tell me about your role. What do you think the role of Ministry of Health and Ministry of Education and Vocational Training in the integrated school eye health programme is currently?  b. What do you think the role of Ministry of Health and Ministry of Education and Vocational Training in the integrated school eye health programme should be?  c. Do you think the existing working relationship works well? Are there gaps? Where the overlaps and synergies than can are exist in the two ministries?  d. What would you suggest for better coordination for a better integrated school eye health programme?  e. Going forward how do you see the programme being financed? Are there any opportunities for budget allocation for integrated school eye health programme? |
| How will the integrated school eye health programme be managed and implemented? | **2. Training specific** | a. What do you think about the training given to the teachers in terms of:   - Time of the training - Training materials - Mode of delivery - Trainers of the training - Duration of the training   b. How do you think we can improve it? |
|  | **3. Vision screening** | a. What do you think about the vision screening for the children?  b. Has is work well? What worked? What did not work? |
| What is the acceptability of the stakeholders towards an integrated school eye health programme? | **4. Acceptability of the interventions to school teachers, communities and students** | a. What do you think about the integrated school eye health programme?  b. What are the benefits?  c. What are the disadvantages? |
|  | **5. Perceptions on the integrated programme** | a. What do you understand about integrated school eye health programme?  b. Tell me something about how the programme and integration could be strengthened and why.  c. Do you think it is easy to combine interventions at the school level? |
| What are the roles that different department plays and how can we share resources with health programmes that have similar interests in eye health? | **6. General comments on the overall programme, including time spent on the integrated programme, and recommendations for streamlining and strengthening.** | a. What so you see as a source of resources for schools to maintain the integrated school eye health programme?   b. How do you think we can combine efforts with eye health? c. Which other health initiatives that the government think will benefit from an integrated school health programme? |
